# Double face of glycolytic enzyme ENO1 in heart failure

**DOI:** 10.1101/2025.03.31.25324995

**Authors:** Shuai Yuan, Rong Xie, Xudong Zhang, Yuyan Tang, Kunying Jin, Jiahui Fan, Chen Chen, Dao Wen Wang, Huaping Li

**Affiliations:** Division of Cardiology, Tongji Hospital, Tongji Medical College, Huazhong University of Science and Technology, Wuhan 430030, China; Hubei Key Laboratory of Genetics and Molecular Mechanisms of Cardiological Disorders, Wuhan 430030, China

**Keywords:** ENO1, heart failure, epigenetics, moonlighting function

## Abstract

**RATIONALE:** Glycolytic enzyme ENO1 is dysregulated in both nucleus and cytoplasm in failing hearts.

**OBJECTIVE:** We aim to elucidate the function and mechanism of nuclear and cytoplasmic ENO1 in pressure-overload induced heart failure.

**METHODS AND RESULTS:** Subcellular fractionation followed by western blotting revealed increase of nuclear and cytoplasmic localized ENO1, a glycolysis enzyme, in transverse aortic constriction (TAC)-induced failing hearts. In vivo study showed that overexpression of nuclear ENO1 exacerbated heart failure while cytoplasmic ENO1 exerted an opposite effect. Mechanistically, cytoplasmic ENO1 activated AKT phosphorylation and increased the contractility of cardiomyocytes. In nucleus, ENO1 bound to NOC2L, a transcriptional repressor, to decrease NOC2L recruitment on glycoprotein nonmetastatic melanoma protein B (GPNMB) promoter, leading to GPNMB transcriptional activation as revealed by immunoprecipitation coupled with mass-spectrometry, cut-tag sequencing and chromatin immunoprecipitation polymerase chain reaction (ChIP-PCR). Furthermore, co-culture assays showed that GPNMB secreted by cardiomyocytes activated cardiac fibroblasts differentiation, leading to augmented pathological cardiac remodeling involving both cardiac hypertrophy and fibrosis through a cardiomyocytes-fibroblast crosstalk.

**CONCLUSIONS:** We uncovered the protective role of cytoplasmic ENO1 and detrimental role of nuclear ENO1 in cardiac pathological remodeling. Inhibition of cardiac nuclear ENO1-GPNMB pathway by rAAV-tnt-shRNA provide new insights for treating heart failure.

## Introduction

Heart failure (HF) refers to a group of clinical syndromes in which ventricular filling and/or ejection function are impaired due to cardiac structural or functional disturbances. Due to the aging population and increased prevalence of obesity, hypertension and diabetes, the incidence of HF will continue to rise^1^. The prognosis of heart failure patients is poor as the 5-year mortality is as high as 25-50%, which brings a heavy burden to the social economy and public health^2, 3^. Therefore, novel mechanisms and therapeutic targets underlying HF are urgently needed.

The widely recognized pathological manifestation of HF is structural remodeling which involves cardiomyocyte hypertrophy, cardiomyocyte apoptosis and fibrosis. However, apart from structural remodeling, other abnormalities are also engaged in HF such as electrophysiological and metabolic remodeling^4^. Multiple studies have shown that metabolic alterations precede other remodeling forms, suggesting that metabolic remodeling might be actively involved in the initiation of HF through metabolites, signaling pathways, and epigenetic regulation^5–8^. In terms of cardiac metabolism, glucose and fatty acids are the main source of energy production for cardiomyocytes. The oxidation of fatty acids provides more than 70% of ATP in cardiomyocytes under physiological conditions while the proportion of energy derive from glucose and lactic acid increases from 30% to about half during myocardial hypertrophy^9, 10^. Although it is widely held that hypertrophied and failing hearts display increased glycolysis^11^, paradoxically, unchanged or decreased glycolysis have also been observed in hypertrophied hearts^12^, which might be due to differences in animal models and/or disease stages^8^. Functionally, a transgenic mouse model expressing kinase-deficient glycolytic enzyme pyruvate kinases type M2 (PKM2) leads to reduced glycolysis, more-profound hypertrophy and cardiac dysfunction in response to pressure overload^13^. However, persistent high rate of glycolysis also induces pathological hypertrophy^14^. Therefore, it seems activated and impaired glycolysis are likewise detrimental and restoration the balance may be beneficial for treating heart failure.

Of note, the rate-limiting enzymes in the glycolytic pathway are hexokinase (HK), phosphofructokinase (PFK), and pyruvate kinase (PKM) while the non-rate-limiting enzymes include phosphoglucose isomerase (GPI), aldolase (ALDOA), GAPDH, phosphoglycerate kinase (PGK), phosphoglycerate mutase (PGAM), enolase (ENO), lactate dehydrogenase (LDH). Glycolytic enzymes are generally considered regulating various metabolic processes in the cytoplasm. However, many studies have revealed their non-canonical function in the nucleus. For example, PKM2 in the cytoplasm promotes the proliferation and repairment of cardiomyocytes after myocardial ischemic injury^15, 16^. Interestingly, PKM2 is also able to enter the nucleus and alleviate the chemotherapy drugs induced-cardiotoxicity by regulating the “switch” of p53^17^. Our group has focused on studying the subcellular functions of RNAs and proteins in HF for decades. We have uncovered distinct roles of the same miRNA or protein (Ago2) in different subcellular fractions^18–21^. Recently, we accidently observed the presence of glycolytic enzyme ENO1 in nucleus, which functions as a lncRNA binding protein to regulate IL6 transcription^22^. However, whether glycolytic enzymes such as ENO1 participate in heart failure and whether they function through canonical or non-canonical manners, remain unclear. In this current study, we try to identify the dysregulated glycolytic enzymes under heart failure and explore the detailed functions and mechanisms.

## Results

### Nuclear ENO1 is up-regulated while the activity of ENO1 is down-regulated during heart failure

To identify dysregulated glycolytic enzymes during heart failure, we examined cardiac expression of glycolytic enzymes in TAC-induced heart failure mice. We separated subcellular fractions and observed that nuclear ENO1 was increased while other enzymes remained largely unchanged or undetectable in nucleus under heart failure (Fig. 1A-B). Moreover, the specific dysregulation of ENO1 (but not other glycolytic enzymes) in nucleus argued against potential cytoplasmic proteins contamination in the isolated nuclear fraction.

**Figure 1.**
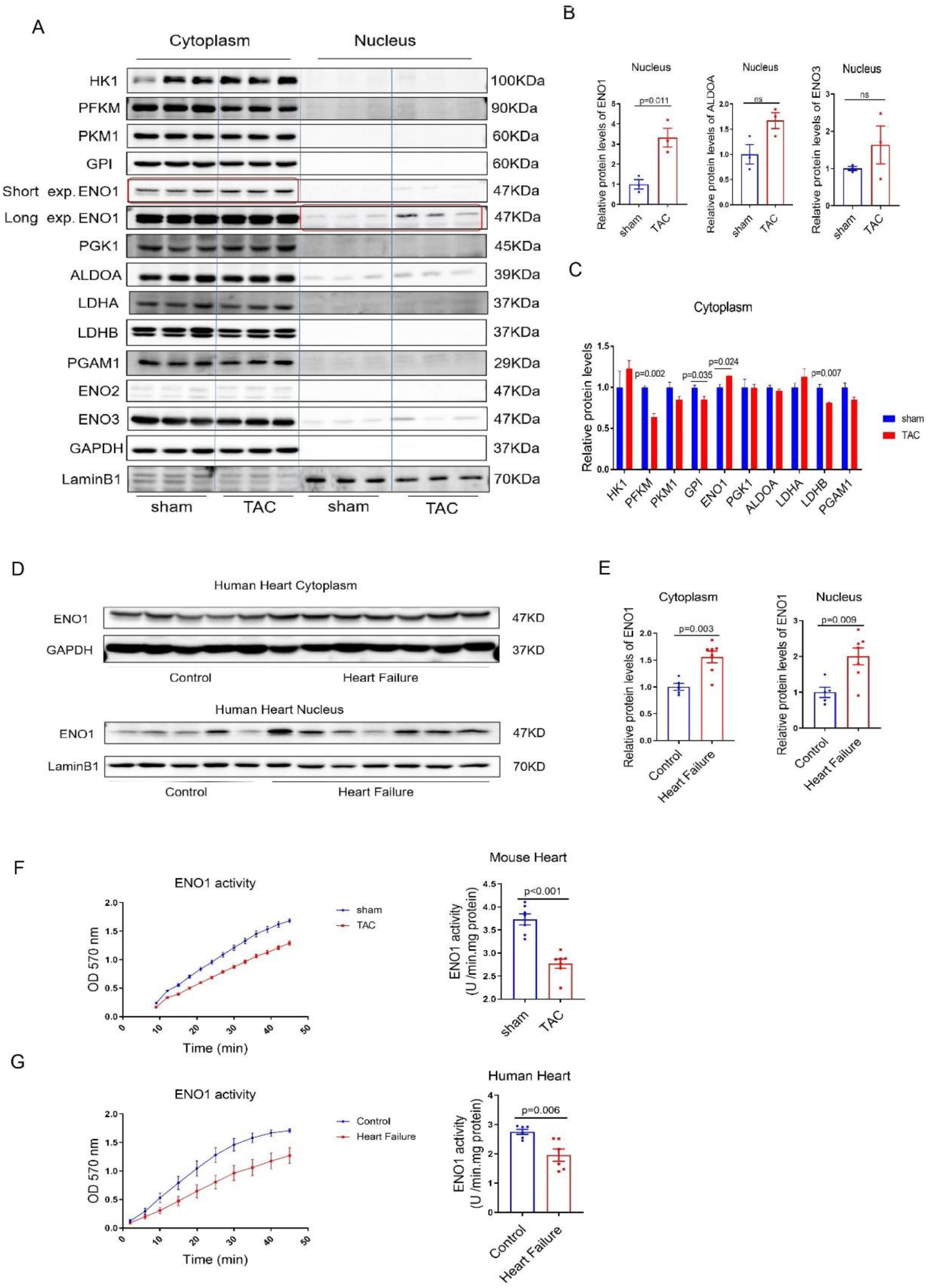
Increased nuclear ENO1 and decreased activity of ENO1 in failing hearts. (A) Western blotting analysis of cytoplasmic and nuclear glycolysis enzymes in mouse hearts treated with sham or transaortic constriction (TAC) operation. (B, C) Statistic analysis of glycolysis enzymes expressions in nucleus (B) and cytoplasm (C). (D, E) Western blotting analysis of ENO1 in cytoplasm (upper) and nucleus (down) in hearts from heart failure (HF) patients or healthy control (D). Statistical analysis of cytoplasm and nuclear ENO1 expression (E). (F) ENO1 enzymatic activity detection of sham or TAC treated mouse hearts. (G) ENO1 enzymatic activity detection of control or HF patients’ hearts. (B, C) N = 3; (E) N = 5 for control, N = 7 for HF; (F) N = 7; (G) N = 6. (B, C, E-G) Student *t* test.

In cytosol, ENO1 was slightly up-regulated while PFKM, GPI, and LDHB were down-regulated in the failing hearts (Fig. 1A-C). We then detected cardiac expression of ENO1 in patients with heart failure, obtaining consistent results that ENO1 expression was increased in nucleus (Fig. 1D-E, Sup. Table 1). Meanwhile, quantitative real-time polymerase chain reaction (qRT-PCR) analysis showed increase of ENO1 mRNA levels in heart failure (Sup. Fig. 1A). Enolase (ENO) has three isoforms, alpha-enolase (ENO1) is ubiquitously present in most tissues, gamma-enolase (ENO2) is mainly expressed in neural tissue and beta-enolase (ENO3) is predominantly expressed in skeletal muscle^23,24^. We proceeded to examine different isoforms of enolase. As a result, other ENOs were not significantly changed in the nucleus under TAC stress (Fig. 1A-B).

Because ENO1 is a glycolytic enzyme, we also analyzed its enzymatic activity, finding down-regulated ENO1 activity in both TAC-treated mice heart and DCM patients despite an increase of its protein level (Fig. 1F-G). In summary, these data revealed that nuclear and cytoplasmic ENO1 were increased while the enzymatic activity of ENO1 appeared to be decreased during heart failure.

### Cytoplasmic ENO1 protects against whereas nuclear ENO1 augments cardiac dysfunction in TAC-induced heart failure mice

According to the published single-cell sequencing data from mice heart^25^, ENO1 mRNA was detectable in cardiomyocytes but not in cardiac fibroblasts, cardiac endothelial cells or cardiac macrophages. Therefore, we reasoned that ENO1 might exert a predominant role in cardiomyocytes. To determine the unique effects of subcellular ENO1, we exogenously expressed ENO1 in cytoplasm and nucleus respectively by utilizing nuclear localization sequence (NLS) and nuclear exporting sequence (NES). Immunofluorescence staining confirmed successful and specific subcellular overexpression of ENO1-Flag in cardiomyocytes by using recombinant adeno-associated virus (rAAV) combined with a cardiac-specific troponin T (TNT) promoter (Fig. 2A, Sup. Fig. 2A). Interestingly, echocardiography data revealed that cytoplasmic ENO1 protected against whereas nuclear ENO1 augmented TAC-induced cardiac dysfunction (Fig. 2B). Further cardiac hemodynamic examination showed consistent regulation patterns as echocardiography (Fig. 2C). In accordance, TAC-induced cardiomyocytes hypertrophy and cardiac fibrosis were ameliorated in cytoplasmic ENO1 overexpressed mice while aggravated by nuclear-ENO1 overexpression (Fig. 2D-G). We further analyzed glycolysis products, finding that lactate and pyruvate were up-regulated by cytoplasmic ENO1 overexpression but not in nuclear ENO1 administrated hearts (Sup. Fig. 2B-C).

**Figure 2.**
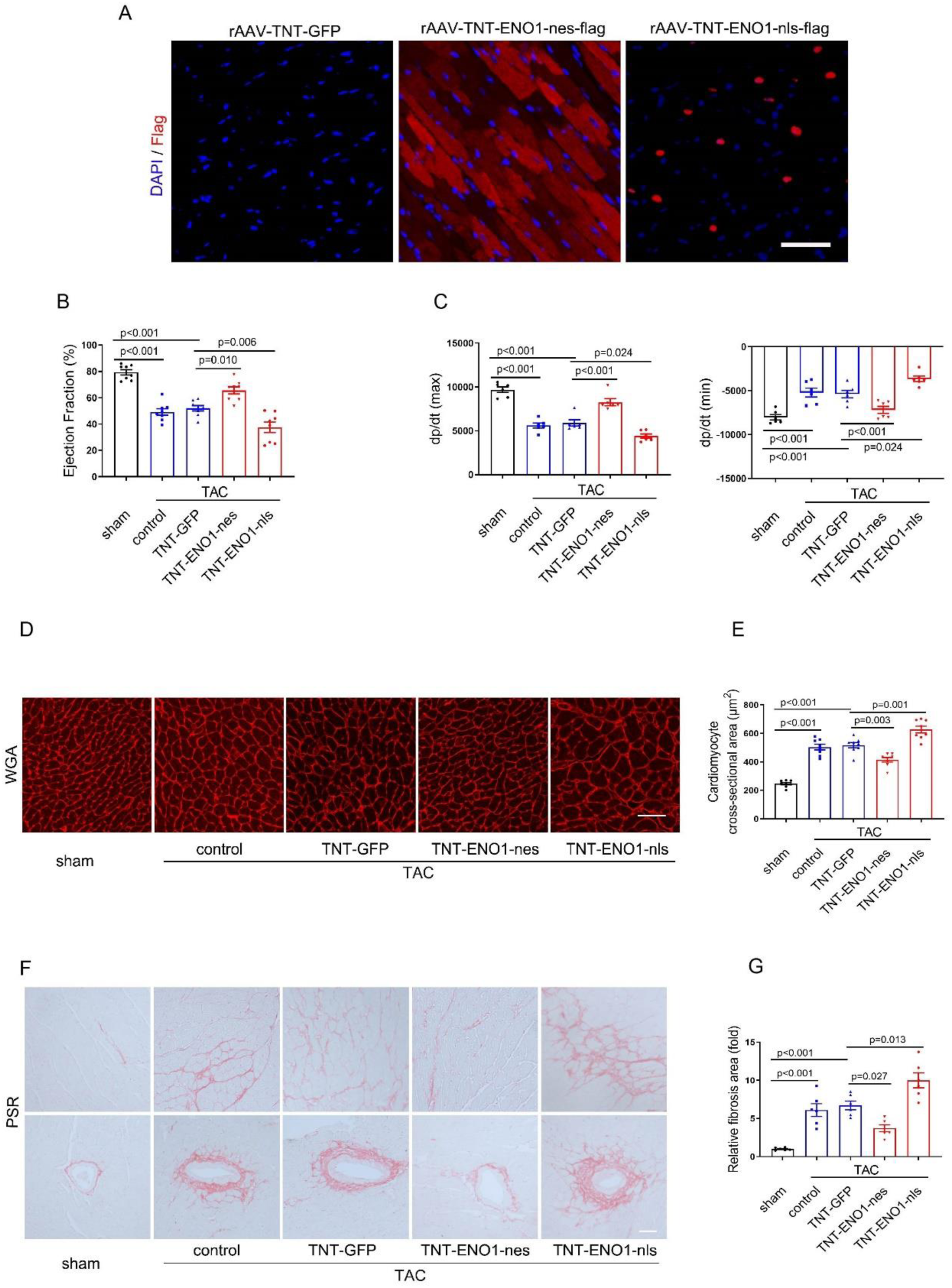
Cytoplasmic ENO1 protected while nuclear ENO1 deteriorated cardiac dysfunction. (A) Flag immunofluorescence staining of heart sections from control (rAAV-TNT-GFP), cytoplasmic ENO1 overexpression (rAAV-TNT-ENO1-nes-flag) and nuclear ENO1 overexpression (rAAV-TNT-ENO1-nls-flag) mouse. Scar bar, 50 μm. (B) Ejection fraction analysis by echocardiograms of indicated groups. (C) Hemodynamic analysis of cytoplasmic or nuclear ENO1 overexpressed mouse. (D, E) WGA staining of cardiac sections in indicated groups. Representative image (D) and statistical analysis (E). Scar bar, 50 μm. (F, G) Picro Sirius Red (PSR) staining of cardiac sections in indicated groups. Representative image (F) and statistical analysis (G). Scar bar, 50 μm. (B, E) N = 8; (C, G) N = 6. (B, C, E, G) One-way ANOVA followed by Tukey’s multiple comparisons test.

Next, we investigated whether knockdown of ENO1 had any regulatory effects on cardiac function by utilizing rAAV-ENO1-shRNA (Sup. Fig. 2D-F). Echocardiography and hemodynamic examination showed no significant differences in EF value and dp/dt between control and ENO1-knockdown group (Sup. Fig. 2G-H). Moreover, knockdown of ENO1 was unable alter TAC-induced cardiac hypertrophy and fibrosis (Sup. Fig. 2I-L).

Taken together, these data suggested that cytoplasmic ENO1 mitigated whereas nuclear ENO1 aggravated cardiac dysfunction in TAC-induced heart failure. However, ENO1 knockdown by shRNA failed to show any detectable changes in cardiac performance, which might be due to a combinational effect mediated by cytoplasmic and nuclear ENO1 knockdown.

### Cytoplasmic ENO1 improves cardiomyocytes contractility via AKT activation

To explore the mechanisms of subcellular ENO1, we overexpressed exogenous ENO1 in cultured AC16 cardiomyocytes by ultilizing nuclear exporting sequence and nuclear localization sequence (Fig. 3A). Previosu studies have shown that ENO1 regulates various biological functions by activating AKT at Thr 308 and Ser 473^26–29^. AKT activation positively regulates contraction by increasing calcium influx^30, 31^. Moreover, loss of AKT profoundly exacerbates TAC-stimulated cardiac hypertrophy^30^. A step further, in this stuy we determined to explore the difference of nuclear and cytoplamic ENO1 regarding AKT regulation. For this end, isoproterenol (ISO) was used to induce NRCM pathological state in which AKT phosphorylation was dramatically inhibited (Sup. Fig. 3A, Fig. 3B-C). A similar downreulation was also observed in TAC pressure-overload hearts (Fig. 3D-E). Interestingly, cytoplasmic ENO1 restored the downregulation of AKT phosphorylation induced by ISO or TAC while nuclear ENO1 had no such effects (Fig. 3B-E). It is well recognized that sarcomere shortening is an indicator for cardiomyocyte contractility^32–34^. Notably, cytoplasmic ENO1 overexpression pronouncedly alleviated pressure overload-induced damage of contractility as indicated by the restore of sarcomere shortening return velocity, departure velocity, and fractional shortening in mouse primary cardiomyocytes (Sup. Fig. 3B, Fig. 3F-H). To further determine whether the upregulation of cytoplasmic ENO1 on contractility was dependent on AKT activation, we treated isoalted adult cardiomyocytes with two different AKT phosphorylation inhibitors (GSK690693, MK2206) respectively. Under these conditions, cytoplasmic ENO1 was no longer able to enhance the reduced sarcomere shortening (Fig. 3I-K).

**Figure 3.**
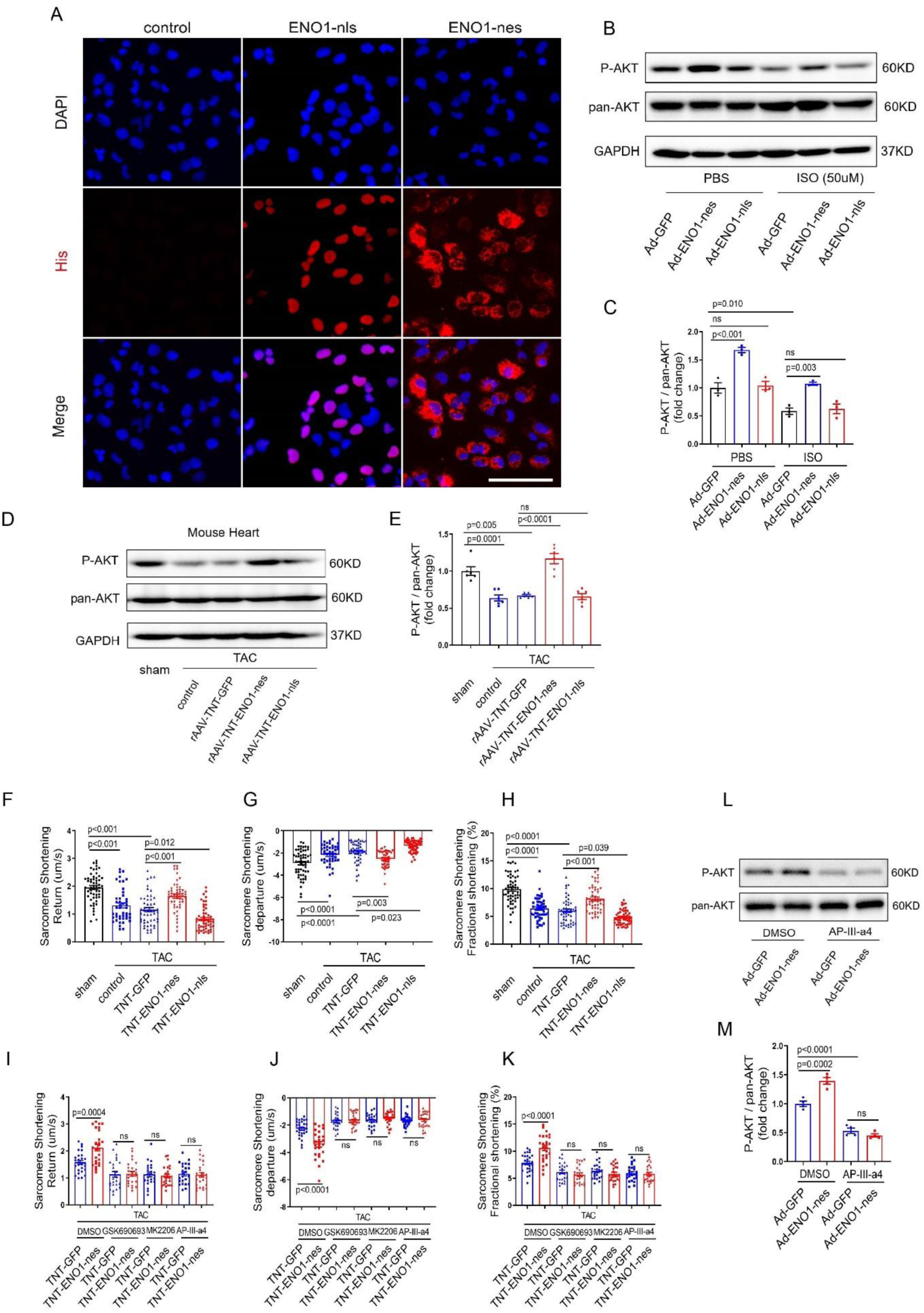
Cytoplasmic ENO1 alleviated cardiomyocytes contractile dysfunction via AKT activation. (A) His immunofluorescence staining of AC16 cardiomyocytes with nuclear ENO1 (ENO1-nls) or cytoplasmic ENO1 (ENO1-nes) overexpression. Scar bar, 100 μm. (B, C) Western blotting detection of P-AKT and pan-AKT in cytoplasmic or nuclear ENO1 overexpression neonatal rat cardiomyocytes (NRCM) treated with isoproterenol (ISO, 50 μM) (B). Statistical analysis of the ratio between P-AKT (473) and pan-AKT (C). (D, E) Western blotting detection of P-AKT (473) and pan-AKT in mouse hearts of indicated groups (D). Statistical analysis of the ratio between P-AKT and pan-AKT (E). (F, G, H) Primary cardiomyocytes contractility analysis indicated by sarcomere shortening return velocity (F), departure velocity (G) and fractional shortening (H) in mice with different treatment. (I, J, K) Primary cardiomyocytes contractility analysis indicated by sarcomere shortening return velocity (I), departure velocity (J) and fractional shortening (K) in cardiomyocytes treated with AKT phosphorylation inhibitor (GSK690693 1 μm, MK2206 1 μm) or ENO1 inhibitor (AP-III-a4 10 μm). (L, M) Western blotting analysis P-AKT (473) and pan-AKT in NRCM (L) and statistical analysis of P-AKT/pan-AKT (M). (C, M) N = 3; (E) N = 6. (C, E-H, M) One-way ANOVA followed by Tukey’s multiple comparisons test. (I-K) Student *t* test.

Since we have observed declined enzyme activity of ENO1 during heart failure (Fig. 1F-G), we then tested whether the regulation of cytoplasmic ENO1 on contractility was dependent on its enzymatic activity. We treated primary cardiomyocytes isolated by perfusion with antagonist of ENO1 (AP-III-a4), observing the up-regulation of contractility by cytoplasmic ENO1 was lost (Fig. 3I-K). In accordance, increased AKT phosphorylation by cytoplasmic ENO1 overexpression was also lost under AP-III-a4 treatment (Fig. 3L-M). Moreover, we found that cytoplasmic ENO1 treatment increased ENO1 enzymatic activity in TAC-operated hearts (Sup. Fig. 3C). Together, these data suggested that cytoplasmic ENO1 overexpression protected against TAC-induced cardiomyocytes contractile dysfunction, which appeared to be meidated by AKT activation through increased enzymatic activity of ENO1.

### NOC2L binds to ENO1 in nucleus of cardiomyocytes

To explore the mechanism of nuclear ENO1, we first identify the protein partners for nuclear ENO1 in cardiomyocytes. As shown in Fig. 4A, nucleus of human AC16 cardiomyocytes was isolated followed by ENO1-immunoprecipitated for mass spectrometry (MS) detection (Sup. Fig. 4A). A total of 18 proteins were specifically captured by ENO1-IP compared to IgG-IP control (Sup Table 2, Sup EXCEL 1). Next, we screened for nucleus-localized proteins, and according to Genecard database, 7 out of 18 proteins were expressed in the nucleus, among which 5 displayed higher difference scores compared to IgG-IP (NACC1, FAU, RBBP6, NOC2L, SAP30BP, Sup Table2).

**Figure 4.**
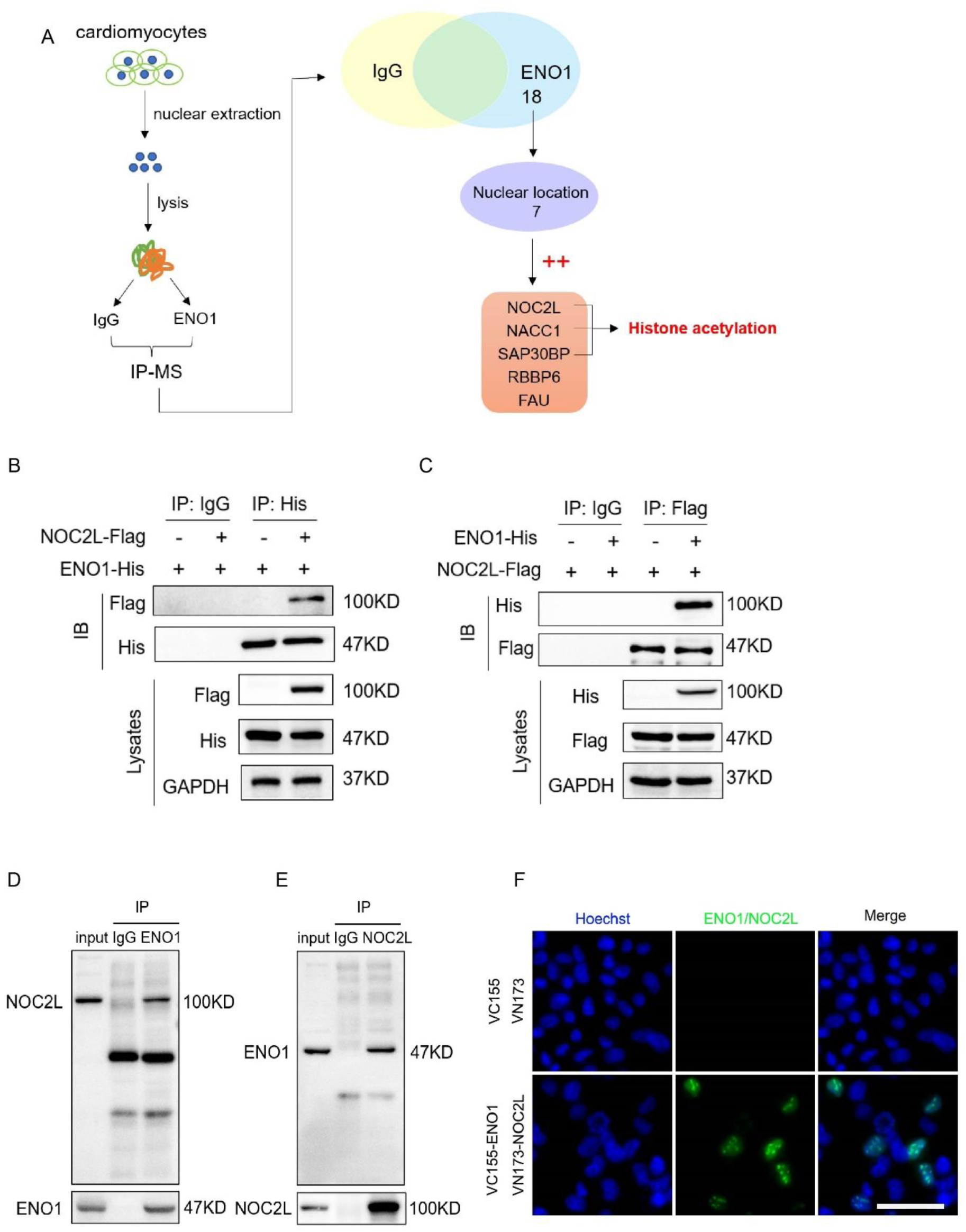
Nuclear ENO1 combined with NOC2L independent of PEP. (A) Schematic diagram of the screening strategy for identifying nuclear ENO1 binding proteins. (B) IP by His-antibody followed by western blotting detection of Flag in 293T cells co-transfected with ENO1-His and NOC2L-flag. (C) IP by Flag-antibody followed by western blotting detection of His in 293T cells co-transfected with ENO1-His and NOC2L-flag. (D) ENO1-IP followed by western blotting detection of NOC2L in AC16 cardiomyocytes. (E) NOC2L-IP followed by western blotting detection of ENO1 in AC16 cardiomyocytes. (F) BiFC assay in AC16 cardiomyocytes transfected with VC155-ENO1 and VN173-NOC2L. Scar bar, 20 μm.

Notably, among the 5 proteins, 3 nucleoproteins (NOC2L, NACC1, SAP30BP) are reportedly relate to histone acetylation modification^35–37^ and acetylation modifications of histones actively participate in cardaic remodeling^38^. We then evaluated the binding capabilityof the aforementioned 3 candidate proteins (NOC2L, NACC1, SAP30BP) with ENO1 by co-immunoprecipitation (co-IP). By exogenous overexpression in 293T cells, ENO1 was able to bind to NOC2L, and reversely, NOC2L-IP sucessfully captured ENO1 (Fig. 4B-C, Sup. Fig. 4B-C). In AC16 cardiomyocytes, endogenous ENO1 and NOC2L were closely bound to each other (Fig. 4D-E). To directly visualize ENO1-OC2L interactions in living cells, we performed bimolecular fluorescence complementation (BiFC) assay^39^, finding that ENO1 and NOC2L were closely associated in the nucleus of AC16 cardiomyocytes (Fig. 4F). Furthermore, to test whether this binding was ENO1 enzymatic activity dependent, we performed IP experiments pre-treated with AP-III-a4, ENO1 enzymatic activity blocker, and 2-Phosphoglycerate (2-P-G), the metabolic substrate of ENO1. As a result, AP-III-a4 or 2-Phosphoglycerate treatment had no effects on ENO1-NOC2L binding event (Sup. Fig. 4D). In terms of ENO1-NOC2L binding, both RNA level and protein level of NOC2L were not regulated by nuclear ENO1 overexpression (Sup. Fig. 4E-F). Together, these results demonstrated that ENO1 bound to NOC2L in the nucleus, which was independent of its enzymatic activity.

### ENO1 sequesters NOC2L to promote GPNMB transcription via enhancing H3K27 acetylation

NOC2L, also called NIR (Novel INHAT Repressor), was able to bind to nucleosomes and histones, blocking their acetylation mediated by histone acetyltransferases, thus acting as a bona fide INHAT (inhibitor of histone acetyltransferases)^35, 40^. To explore the direct target of ENO1/NOC2L complex, we performed mRNA sequencing in nuclear ENO1 overexpressed NRCM. A total of 950 genes (fold change > 2, adj.P. > 0.05, 432 increased and 518 decreased) were changed (Sup Excel 2). To screen out genes with pathological significance, we also performed mRNA-sequencing in TAC-induced failing hearts, and a total of 535 genes (fold change > 2, adj.P. > 0.05, 356 increased and 179 decreased) were changed (Sup Excel 3), in which 62 genes were overlapped with nuclear ENO1 regulated genes (Fig. 5A). QPCR validation of the top 20 genes according to fold change revealed 9 out 20 genes were steadily regulated by nuclear ENO1 in NRCM (Fig. 5B). Because these genes were changed at mRNA levels, we performed cut-tag sequencing of H3K27Ac, a transcriptional activation marker to evaluate chromatin status of these genes. In NRCM, H3K27ac peaks were observed on promoters of GPNMB, TGFB3, SGK1, DUSP2, and ALDH1A3 (Fig. 5C, Sup. Fig. 5A). However, nuclear ENO1 overexpression only altered H3K27Ac occupancy on GPNMB promoter, indicating transcriptional regulation of GPNMB by nuclear ENO1 treatment. We then evaluated GPNMB protein level in cultured cardiomyocytes (HL-1 cells) as well as in mouse hearts, finding consistent up-regulation of GPNMB by nuclear ENO1 overexpression (Fig. 5D, E, Sup. Fig. 5B, C). In terms of other proteins such as TGFB3, ALDH1A3, DUSP2, they showed different regulation patterns in rat NRCM, mouse HL-1 and mouse hearts, and were not studied further.

**Figure 5.**
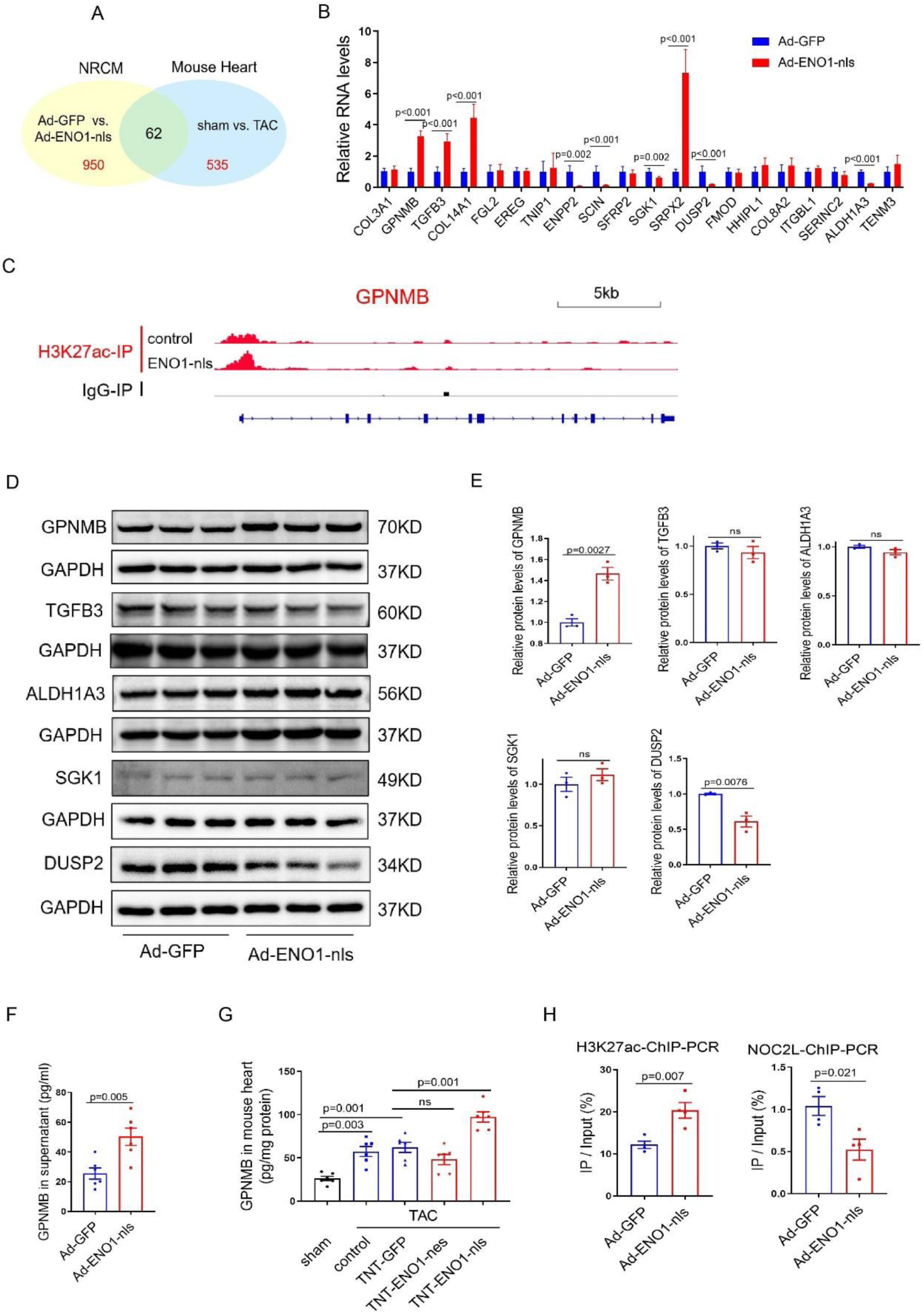
Nuclear ENO1 eliminated the suppression of NOC2L on H3K27ac and promoted GPNMB transcription. (A) Venn diagram of nuclear ENO1 regulated genes in NRCM and TAC induced dysregulated genes in mouse heart. (B) QPCR analysis of the most significantly changed 20 genes in NRCM among the intersection in figure 5A. (C) IGV scan of NRCM cut-tag sequencing data across the GPNMB gene loci. (D, E) Western blotting analysis of GPNMB, TGFB3, ALDH1A3, SGK1, DUSP2 in HL-1 cells treated with Ad-GFP or Ad-ENO1-nls (D). Statistical analysis of the image (E). (F) ELISA analysis of GPNMB in supernatant of AC16 cardiomyocytes with different treatment. (G) ELISA analysis of GPNMB in mouse heart homogenate with different treatment. (H) H3K27ac and NOC2L ChIP-PCR in AC16 cardiomyocytes of GPNMB promoter. (B, F, G) N = 6; (E) N = 3; (H) N =4. (B, E, F, H) Student *t* test. (G) One-way ANOVA followed by Tukey’s multiple comparisons test.

GPNMB, also known as dendritic cell–heparin integrin ligand and Osteoactivin, is a type I membrane glycoprotein consisting of an N-terminal signal peptide, a transmembrane helix and a short C-terminal cytoplasmic domain^41^. Because of the ability of GPNMB to secretion into extracellular space, we detected GPNMB protein level in the supernatant of cultured AC16 and HL-1 cardiomyocytes by ELISA, observing increased GPNMB secretion by nuclear ENO1 treatment (Fig. 5F, Sup. Fig. 5D). Besides, GPNMB was also elevated in the hearts and plasma of mice overexpressing ENO1 specifically in the nucleus (Fig. 5G, Sup Fig. 5E).

Mechanistically, we proposed two possible models to elucidate the regulation pattern of ENO1/NOC2L complex on downstream genes (Sup. Fig. 5F). The upper schematic diagram depicted that ENO1 recruited NOC2L to the promoter region followed by inhibition of H3K27ac and therefore decreased expression of downstream genes. In contrast, the alternative diagram depicted that ENO1 played a sequestering role by binding NOC2L, a transcriptinal supressor, to decrease NOC2L binding on gene promoter. To identify which model is the case for ENO1/NOC2L regulation on GPNMB transcription, we scanned the GPNMB promoter region by genome browser viewer, finding that nuclear ENO1 overexpression led to enhanced H3K27ac signal around GPNMB promoter (Fig. 5C), which suggesed the second diagram was the case. Furthermore, ChIP-PCR revealed no combination between ENO1 and GPNMB promoter (Sup. Fig. 5G). Moreover, ChIP-PCR revealed activated H3K27ac but inhibited NOC2L binding on GPNMB promoter by nuclear ENO1 overexpression in AC16 and HL-1 cardiomyocytes (Fig. 5H, Sup. Fig.5H, I). Furthermore, ENO1 enzymatic blocker AP-III-a4 had no effects in regulating H3K27ac or NOC2L binding events on GPNMB promoter in AC16 cardiomyocytes (Sup. Fig. 5J). In summary, nuclear ENO1 binds to NOC2L and inhibits its ability to target on GPNMB promoter, leading to activated GPNMB transcription.

### Nuclear ENO1 promotes cardiac pathological remodeling via enhanced secretion of GPNMB

To explore whether nuclear ENO1 exacerbated cardiac dysfunction through GPNMB-dependent manner in vivo, we injected rAAV-tnt-sh-GPNMB into TAC-treated mice to knockdown GPNMB expression (Sup. Fig. 6A-C). We noticed that nuclear ENO1 mediated cardiac-dysfunction aggravation was abolished by GPNMB knockdown (Fig. 6A, Sup. Fig. 6C). Consistently, coordinated changes in the size of cardiomyocytes and fibrosis were also observed (Fig. 6B-E), indicating that nuclear ENO1 augmented cardaic remodeling through GPNMB dependent manner. Mechanistically, previous study has reported that GPNMB positively regualtes ANP, BNP and MYH7 expression^42^. We analzyed animal hearts, finding that cardiac GPNMB knockdown abolished nuclear ENO1 induced increase of ANP, BNP and MYH7 (Fig. 6F). According to literature, overexpressed MYH7 was able to impair cardiomyocytes systolic function and accelerate TAC-induced cardiac hypertrophy^43, 44^.

**Figure 6.**
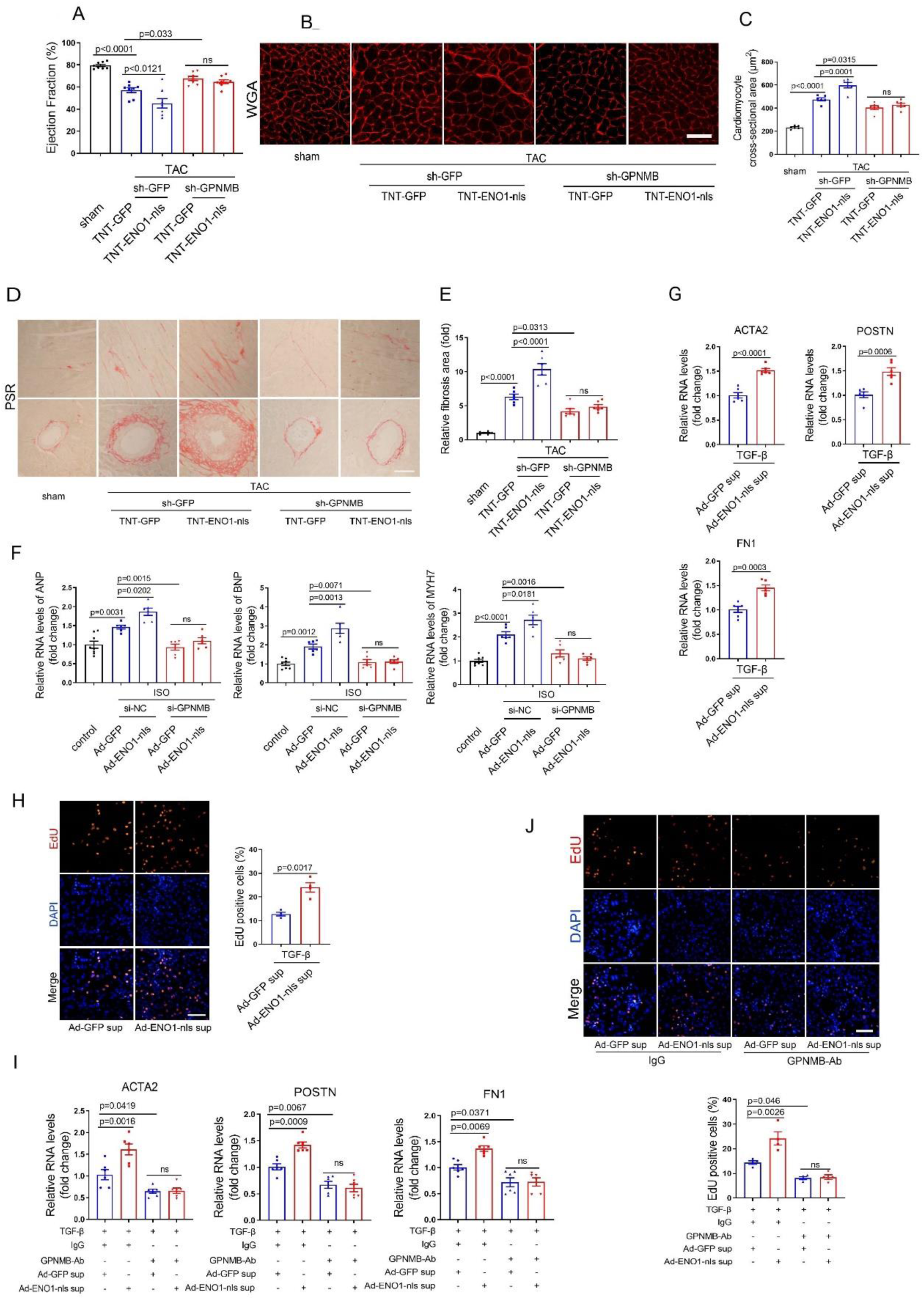
Nuclear ENO1 augmented cardiac dysfunction in dependent of GPNMB. (A) Ejection fraction analysis by echocardiogram of mouse with different treatment. (B, C) WGA staining of cardiac sections in indicated groups. Representative image (B) and statistical analysis (C). Scar bar, 50 μm. (D, E) PSR staining of cardiac sections in indicated groups. Representative image (D) and statistical analysis (E). Scar bar, 200 μm. (F) QPCR analysis of ANP, BNP, MYH7 in HL-1 cells of indicated groups. (G) QPCR analysis of ACTA2, POSTN and FN1 of mouse cardiac fibroblasts treated with cardiomyocytes culture supernatants. (H) EdU analysis of mouse cardiac fibroblasts treated with cardiomyocytes culture supernatants. Representative image (left) and statistic diagram (right). Scar bar, 50 μm. (I) QPCR analysis of ACTA2, POSTN and FN1 of mouse cardiac fibroblasts treated with cardiomyocytes culture supernatants and GPNMB neutralization antibody or control. (J) EdU analysis of mouse cardiac fibroblasts treated with cardiomyocytes culture supernatants and GPNMB neutralization antibody or control. Representative image (upper) and statistic diagram (down). Scar bar, 50 μm. (A, C) N = 8; (E-G, I) N = 6; (H, J) N = 4. (A, C, E, F, I, J) One-way ANOVA followed by Tukey’s multiple comparisons test. (G, H) Student *t* test.

Apart from cardiac hypertrophy, animal data showed that fibrosis was also changed, we thus asked how nuclear ENO1 overexpression in cardiomyocytes by rAAV-tnt-vector activated fibrosis. To understand the crosstlk between cardiomyocytes and fibroblasts, we performed indirect co-culture experiments. Fibroblasts (NMCF) were cultured with conditioned supernatant isolated from cardiomyocytes (HL-1) as depicted in Sup. Fig. 6D. Under TGF-β treatment, a well established fibrosis activator, nuclear ENO1 pre-treated cardiomyocytes derived supernatant dramatically increased fibroblasts activation markers including ACTA2, POSTN and FN (Fig. 6G). Moreover, EdU assay demonstrated that ENO1-nls overexpressed cardiomyocytes derived supernatant promoted fibroblasts proliferation significantly (Fig. 6H). Next, we performed co-culture assay with neutralizing GPNMB antibody (Sup. Fig. 6E). Interestingly, supernatant from ENO1-nls overexpressed cardiomyocytes pre-treated with GPNMB antibody failed to promote the activation and proliferation of fibroblasts (Fig. 6I, J). These data indicated that nuclear ENO1 induced GPNMB was able to translocate from cardiomyocytes to fibroblasts to promote fibroblasts proliferation and differentiation.

Collectively, these data revealed distinct mechanisms of subcellular ENO1 in cardiac remodeling. Cytoplasmic ENO1 promoted AKT phosphorylation, protected against cardiac dysfunction, whereas nuclear ENO1 sequestered NOC2L function via moonlighting capability. Decreased NOC2L binding on GPNMB promoter enhanced GPNMB transcription. GPNMB mediated cardiac pathological remodeling through cardiomyocytes-fibroblasts crosstalk manner (Fig. 7).

**Figure 7.**
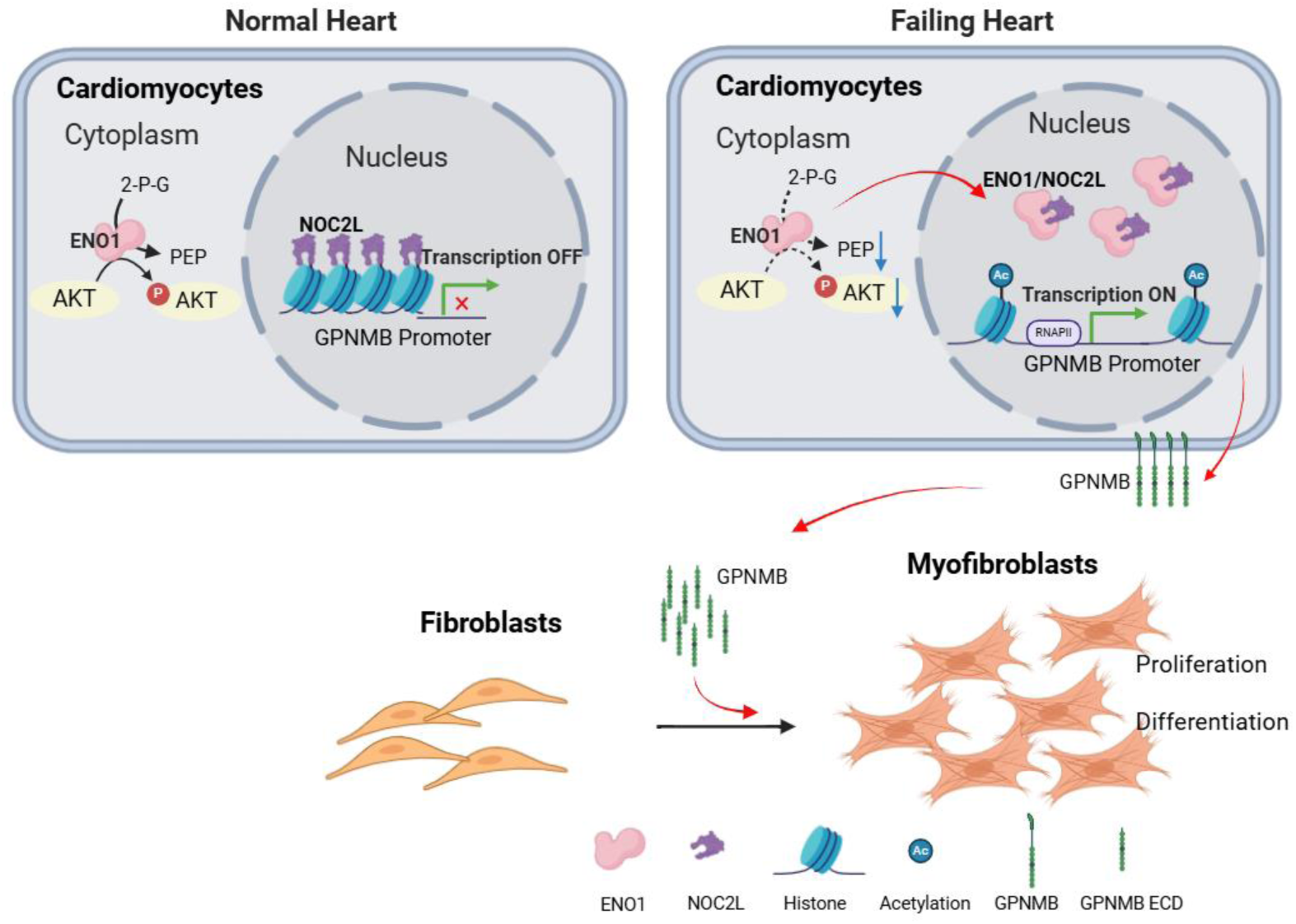
Mechanism pattern diagram of nuclear ENO1. In the normal heart, NOC2L blocked the histone acetylation sites leading to low acetylation of histones in the GPNMB promoter region and condensed chromosome. The transcription of GPNMB was inhibited. In the failing heart, ENO1 translocated into the nucleus and bound to NOC2L, exposing histone acetylation sites in the GPNMB promoter region, enhancing acetylation levels, increasing GPNMB transcription, and further secreting into the extracellular, which aggravating the proliferation and differentiation of cardiac fibroblasts.

## Discussion

Our results demonstrated that a glycolysis enzyme, ENO1 was upregulated in both cytoplasm and nucleus of failing hearts. Strikingly, cytoplasmic ENO1 exerted cardioprotective role whereas nucleic ENO1 aggravated HF by promoting the expression of GPNMB. Importantly, we show that cardiac GPNMB knockdown by rAAV-tnt-shRNA was able to effectively rescue cardiac dysfunction in TAC-induced HF, which provides a new drug therapeutic target for HF.

### Decreased enzymatic activity of cytosl ENO1 in heart failure

Recent studies reported that metabolic remodeling preceds most other changes such as structural remodeling during heart failure, which may play a pathogenic causal role ^45, 46^. The adult heart mainly metabolizes fatty acids to produce energy, while the oxidation of glucose and lactate is relatively small. However, under conditions of hypertrophy, the proportion of glucose utilization in ATP production significantly increases^47^. Compared to the control group, the contribution of glycolysis to energy production of hypertrophic hearts increased ∼2-3 fold^48, 49^. Previous study revealed that glycolytic enzyme LDHA promoted cardiac adaptive growth through elevation of NDRG3 and activation of ERK^10^. Another glycolytic enzyme PKM2 regulated cardiomyocyte cell cycle and promoted cardiac regeneration^15^. ENO1 has been widely investigated in caners. Fu et al. found ENO1 promoted non-small cell lung cancer cells glycolysis, growth, migration, and invasion through FAK-mediated PI3K/AKT pathway^27^. Dai et al. reported that ENO1 regulated the malignant phenotype of pulmonary artery smooth muscle cells via the AMPK-Akt pathway^26^. Inhibition of ENO1 was reported to antagonize the proliferation of cardaic fibrobalsts^50^. Howerer, the subcellular role of ENO1 in heart failure is unclear. In this study, we demonstrated that overexpression of cytoplamic ENO1 improved cardiac function in TAC induced heart failure. Mechanistially, in vitro studies showed that the protective effects of cytoplasmic ENO1 on myocardial contractility were dependent on AKT activation. Previous studies reported that adenovirus-mediated AKT overexpression alleviated cardiomyocyte apoptosis and limited infarct size following ischaemia/reperfusion injury^51, 52^. Moreover, AKT appears to enhance cardiomyocytes contraction by increasing calcium influx through the LTCC^53^, by increasing SERCA2a protein levels^54^, and by augmenting PLB phosphorylation^55^. Therefore, AKT activation might be one of the links between increased ENO1 enzymatic activity and increased cardaic performance. However, it was reported that loss of AKT or long-term over-activated AKT both resulted in cardaic hypertrophy and cardiac dysfunction^30,31^, therefore, targeting ENO1-AKT pathway for treating HF should be cautious about potential dosage- or time-depended detrimental effects.

In this study, we observed decrease of ENO1 enzymatic activity in failing hearts despite an increase of ENO1 protein level. Previous work suggested that ENO1 S282 phosphorylation was responsible for decreased ENO1 activity in HCT116 cells^56^. Whether this is also the case for decresed ENO1 activity in the failing heart and whether other post-translational modifications (PTMs) are involved are interesing subjects for future research. Nevertheless, our data showed that cytoplasmic ENO1 overexpression improved ENO1 enzymatic activity, which might be due to incresed ENO1 moleculars as ENO1 enzymatic activity measured in this study was normalized to a certain amont of total cardaic proteins. Whether the activity of each exogenous overexpressed ENO1 molecular was changed remained unclear.

Another issue rearding ENO1 enzymatic activity is glycolysis seems to be upregulated in hypertrophy heart according to previous studies ^57, 58^, which is contradictory to decreased ENO1 enzymatic activity we observed in the failing hearts. We reason that ENO1 enzymatic activity is not necessarily eaqul to glycolysis rate because glycolysis is also determined by the activity of other glycolytic enzymes. Moreover, other enolases might also play important roles, such as ENO3 which according to single-cell sequencing data has much higher abundance in cardiomyocytes than ENO1. The enzymatic activities of ENO2 and ENO3 in regulation of glycolysis, cardiac hypertrophy and heart failure require further investigation.

### Nuclear ENO1 in transcription control

Previously studies have reported that MBP-1, a 37 kDa protein alternatively translated from the full-length ENO1 mRNA, preferentially localized in the COS-7 cells nuclei and bound to the c-myc promoter, acting as a transcriptional repressor^59^. However, MBP-1 was undetetable in heart according to our previous study^22^, in which ENO1 antibody was capable of recognizing both ENO1 and MBP-1^60^. Therefore, MBP-1 and ENO1 expression appears to be tissue specific. Apart from working with lncRNAs to activate gene transcription^22^, in this study, we show ENO1 is also able to act as “decoy” to sequester certain transcritional regulators, such as NOC2L, which function as a bona fide inhibitor of histone acetyltransferases by locking the acetylation site, to indicrectly acitivate transcription. Whether nuclear ENO1 functions directly as a transcritional factor or indirectly as a sequester for other transcritional factors/co-factors might depend on partner lncRNAs or TFs that are cell-type specific or disease-specific expressed.

However, the specific molecular mechanism by which ENO1 translocate into the nucleus is currently unclear. In fact, many glycolytic enzymes located in the cytoplasm have the ability to translocate to non-cytoplasmic fractions to exert non-metabolic functions. For example, phosphorylated PGK1 was able to translocate to mitochondria to activates pyruvate dehydrogenase kinase isozyme 1 (PDHK1) to inhibit pyruvate metabolism, thereby enhancing aerobic glycolysis^61^. Moreover, PGK1 can also translocate into nucleus to alleviates ADP-dependent inhibition of CDC7, thus promoting DNA replication^62^. Further studies such as post-translational modifications-mass spectrum (PTM-MS) may shed light on ENO1 subcellualr localization mechanisms.

Apart from translocating into nucleus, several studies reported that ENO1 was also expressed on the surfaces of monocytes and macrophages, acting as a novel stimulatory receptor in inflammation regulation^63, 64^. There are reports showing the presence of ENO1 antibodies in the serum of systemic sclerosis patients, which could induce vascular smooth muscle cell contraction^65, 66^. Therefore, the role of membane ENO1 in HF are interesting subject to study in the future.

### GPNMB mediates the crosstalk between fibroblast and cardiomyocyte

Because of the paradoxical roles of cytoplasmic ENO1 and nuclear ENO1, ENO1 knockdown by shRNA failed to show any detectable changes in cardiac performance, therefore, targeting downstram genes of nulcear ENO1, in this case GPNMB, might be a feasible choice for treating HF. GPNMB is a glycoprotein that consists of a large extracellular domain (ECD) and a short 53 amino acid cytoplasmic tail, connected by a single pass transmembrane domain^67^. The ECD can be shed from the plasma membrane by ADAM10, which then act as a paracrine factor to activate multiple signaling pathways in a variety of cells^41, 68, 69^. Previous study revealed that GPNMB aggravated left ventricular remodeling by promoting matrix metalloproteinase 9 expression in myocardial infarction^42^. In our study, co-culture assays proved that GPNMB produced by cardiomyocytes promoted cardiac fibroblasts proliferation and differentiation. Actually, GPNMB is most enriched in macrophages. The regulatory role of GPNMB in inflammation has been controversial in previous studies, which may explained by inherent differences in disease models used^42, 70, 71^. Furthermore, GPNMB in fibrotic tissue remodeling is not yet conclusively clarified. Bone-marrow macrophage-derived GPNMB reduces fibroblast activation in post-MI cardiac repair^72^. Studies of skeletal muscle and liver injury in mice have indicated GPNMB as an antifibrotic factor, while opposite results exist^73–75^. In our study, GPNMB knockdown in cardiomyocytes by rAAV-tnt-shRNA improved cardiac function. However, whether global GPNMB knockdown by siRNA or ASO or antibody would show cardio-protective effects remain unclear. Additionally, GPNMB might exert distinct functions with different dosage or at different disease stages. Moreover, whether there is any functional differneces between cleaved GPNMB (GPNMB-ECD) and full-length GPNMB also awaits further investigation.

## Methods

### Ethics Statement

Human heart samples of dilated cardiomyopathy were collected from Tongji Hospital (Wuhan, China) between January 2012 and October 2014. Control hearts of normal humans were collected from victims of traffic accidents. The characteristics and medical history of patients and control were listed in Sup Table 1. This study was approved by the Ethics Review Board of Tongji Hospital and Tongji Medical College and conformed to the principles outlined in the Declaration of Helsinki. Written informed consents were obtained from individual subjects or their immediate family members. All animal experiments in this study were approved by the Committee on the Ethics of Animal Experiments of the Animal Research Committee of Tongji College and were carried out in accordance with the recommendations of the Guide for the Care and Use of Laboratory Animals of the National Institutes of Health.

### Cell culture and treatments

DMEM high glucose culture medium and fetal bovine serum (FBS) were purchased from GIBCO (Life Technologies Corporation, Carlsbad, CA). Neonatal rat cardiomyocytes (NRCM) were cultured in DMEM supplemented with 10% FBS and penicillin/streptomycin solution. AC16 cells, 293T cells and NIH3T3 cells were purchased from ATCC and routinely maintained in DMEM with 10% FBS. HL-1 cells were cultured using Claycomb medium (Sigma-Aldrich, 51800C) supplemented with 4 mM L-glutamine,100 μM norepinephrine, and 10% FBS. All cells were maintained at 37 °C in an atmosphere of 5% CO_2_.

Adenovirus (Ad-GFP, Ad-ENO1-nes and Ad-ENO1-nls) were purchased from WZ Biosciences Inc. (Jinan, China). Plasmids were constructed at TIANYI HUIYUAN Co., Ltd (Wuhan, China). Cells were transfected with plasmids according to the manufacturer’s protocol of Lipofectamine 2000 (Invitrogen, Carlsbad, CA). TGF-β were purchased from MedChemExpress, NJ, USA. Neutralizing mouse GPNMB Ab (AF2330) and control mouse IgG1 (MAB002) were purchased from R&D Systems. For indirect co-culture assay, 6h after Ad-GFP or Ad-ENO1-nls treatment, cardiomyocytes were subjected to medium change followed by 10 ng/ml TGF-β treatment for additional 48h. Then GPNMB-neutralizing antibody or control antibody were applied to cardiomyocytes and supernatants were collected for fibroblasts culture for another 48h.

### Primary cardiomyocytes isolation

Primary adult mouse cardiomyocytes were isolated from mice by Langendorff system ^21^. Firstly, hearts were rinsed with perfusion buffer (Tyrode Solution containing 10 mM Taurine and 10 mM BDM) for 5 min, followed by digestion buffer (perfusion buffer with 0.06% collagenase II) for an additional 20 min. Next, hearts were dissected into small pieces in BSA solution and filtered through a 100-um cell strainer. The filtrate was centrifuged at 50 g for 3 min to pellet the cardiomyocytes.

Neonatal rat cardiomyocytes (NRCM) were extracted from rat newborn mice less than 3 days old. Briefly, suckling rat hearts were flushed with D-Hank’s solution and cut into small pieces on ice with scissors, followed by digestion with 0.075% collagenase type 2 (Worthington) in 37°C for 5 minutes. The digested supernatant was added to the serum for neutralization, and undigested tissue pieces continued to be digested. Repeat the process of digestion and neutralization until digestion is complete. The neutralized liquid was filtered with a 70um cell strainer followed by centrifuged at 1000g for 3 minutes. Finally, NRCM were purified by culturing with DMEM in incubator for 2 hours to remove fibroblasts.

### RNA extraction and quantitative RT-PCR

Total RNA was extracted using Trizol (Invitrogen, Carlsbad, CA) and reverse transcription to cDNA by HiScript II 1st Strand cDNA Synthesis Kit (R212, Vazyme, Nanjing, China). The expression of mRNA was quantified by real-time PCR using Power SYBR Green PCR Master Mix (Invitrogen, Carlsbad, CA) on 7900HT FAST real-time PCR system (Life Technologies, Carlsbad, CA). Primers used in the present study were listed in Sup Table 3.

### Subcellular fractions separation

Cytoplasmic and nuclear fractions were separated using the Cell Fractionation kit (Cat. #9038, Cell Signaling Technology, Danvers, MA) according to the manufacturer’s protocol. Western blotting assay was performed to determine cytoplasmic marker GAPDH and nuclear marker laminB1.

### Detection of ENO1 activity

ENO activity in heart tissue or cardiomyocytes was measured using the Enolase Activity Kit (K691, Biovision, Milpitas, CA) according to the manufacturer’s protocol.

### Liquid chromatography-mass spectrometry

Nuclear fraction of AC16 cells was extracted followed by ENO1-immunoprecipitation. The enriched proteins were subjected to mass spectrometry by SpecAlly Life Technology Co., Ltd, Wuhan, China.

### Cell labeling with 5-ethynyl-2’-deoxyuridine (EdU)

Proliferating cells are labeled with EdU and visualized by staining via Cell-Light EdU Apollo567 In Vitro Kit according to the manufacturer’s protocol (C10310, RiboBio Co., Ltd., Guangzhou, China).

### Protein extraction and western blotting

Protein concentrations were measured using the bicinchoninic acid (BCA) method. Cell lysates were mixed with loading buffer and heated at 95°C for 5 minutes. Proteins of different molecular weights were separated by polyacrylamide gel, followed by transferred to nitrocellulose membrane and blocked with Tris-Buffered Saline Tween-20 (TBS-T) containing 5% BSA for 2 hours at room temperature. Membranes were incubated with the indicated primary antibodies overnight at 4°C, followed by incubation with peroxidase-conjugated secondary antibodies for 2 h at room temperature. Finally, protein bands were visualized using an ECL system (Beyotime Institute of Biotechnology, Nanjing, China). Antibodies used in this study were listed in Sup Table 4. Western blot results were densitometry and quantified by Image J software (National Institutes of Health software).

### Transcriptome sequencing

mRNA-seq and data analyses were performed by SeqHealth Biotechnology Co., Ltd (Wuhan, China).

### Chromatin immunoprecipitation (ChIP)-PCR

Cardiomyocytes were crosslinked with 1% formaldehyde for 15 min at room temperature followed by neutralized with glycine for 5 min. Cells washed with PBS were homogenized in lysis buffer and sonicated to generate chromatin fragments with an average size of 200-1000 bp. Fragments were incubated with the indicated antibody or negative control IgG overnight at 4°C. Then the mix were added with Pierce™ Protein A/G Magnetic Beads (Cat# 88802; Thermo Fisher Scientific, Waltham, MA) and mixed by gentle vortex at 4°C for 2 hours. After serial washing, the immunoprecipitated DNA was eluted and purified with a PCR purification kit (Qiagen GmbH, Hilden, Germany). PCR was then performed using primers targeting the promoter region of the selected genes.

### Cut-tag sequencing

Cut-tag sequencing of NRCM to determine the region of indicated proteins or modifications on genome was performed according to the manufacturer’s protocol (TD903, Vazyme, Nanjing, China).

### Enzyme-linked immunosorbent

The concentration of GPNMB in cell culture supernatant, plasma and tissue homogenate were measured according to the manufacturer’s protocol (DY2550, DY2330, R&D).

### Animals

Male C57BL/6 mice aged 8 weeks were purchased from GemPharmatech Co., Ltd (Nanjing, China). Transverse aortic constriction (TAC) was applied to induce pressure overload–induced heart failure. Briefly, mice were anesthetized with pentobarbital (60 mg/kg, intraperitoneal injection), fixed on the operating table, and mechanically ventilated after tracheal intubation with a tidal volume of 0.2 mL and a respiratory rate of 100 times/min. Firstly, the aortic arch was exposed by blunt dissection of the second intercostal space. Then, a 7-0 suture was banded against a 27G needle around the aortic arch. Finally, the needle was pulled out carefully followed by suture of the muscle and skin layer by layer to close the chest cavity with 4-0 silk suture. Sham-operated mice underwent a similar surgical procedure without aortic coarctation. Eight weeks after surgery, all animals were deeply anesthetized by intraperitoneal injection of sodium pentobarbital (60 mg/kg) and sacrificed by exsanguination (part of the descending abdominal aorta). Tissue samples were obtained and frozen in liquid nitrogen, then stored at -80°C.

Construction of plasmids and preparation of recombinant adeno associated virus (rAAV) was supplied by WZ Biosciences Inc. 1 × 10^11^ vector copy numbers of type9 rAAV was given via tail vein to realize cardiac specifically expression. To explore the function of ENO1 in vivo, mice of ENO1-overexpression were randomly divided into 5 groups as follows: sham, TAC, TAC + rAAV-TNT-GFP, TAC + rAAV-TNT-ENO1-nes, TAC + rAAV-TNT-ENO1-nls (n = 8/group) while mice of ENO1 knockdown were randomly divided into 4 groups as follows: sham, TAC, TAC + rAAV-TNT-sh-GFP, TAC + rAAV-TNT-sh-ENO1 (n = 8/group). To evaluate whether the effects of nuclear ENO1 were GPNMB-dependent, male C57BL/6 mice aged 8 weeks were randomly divided into the following 5 groups: sham, TAC + rAAV-TNT-GFP + rAAV-TNT-sh-GFP, TAC + rAAV-TNT-ENO1-nls + rAAV-TNT-sh-GFP, TAC + rAAV-TNT-GFP + rAAV-TNT-sh-GPNMB, TAC + rAAV-TNT-ENO1-nls + rAAV-TNT-sh-GPNMB.

### Cardiac function determination

After induction of anesthesia, echocardiography was performed using a 30-MHz high-frequency Vevo770 scan head (VisualSonics Inc., Toronto, Canada) as described previously ^76^. Left ventricular catheterization was performed to monitor hemodynamics by inserting a Millar 1.4F, SPR 835 catheter manometer (Millar Inc., Houston, TX) into the left ventricle via the right carotid artery. After stabilization, data was recorded continuously. Cardiac function parameters were calculated using LabChart software (Millar Instruments, Inc. Houston, TX) as described ^21^.

### Histologic analysis

Formalin-fixed hearts were embedded in paraffin, sliced into 4-6μm sections and stained with Wheat Germ Agglutinin (WGA) to determine cardiac morphology. Picro Sirius Red (PSR) staining was performed to determine cardiac fibrosis. The tissue sections were visualized by microscope and quantified by Image-Pro Plus Version 6.0 (Media Cybernetics lnc., Bethesda, MD).

### Statistical analysis

Statistical significance was determined as values with a probability (P) of < 0.05 using Prism version 8.0.2 software (GraphPad Software, San Diego, CA). Normality was typically assessed in all datasets regardless of size, using Shapiro-Wilk tests, followed by parametric or nonparametric tests as appropriate. Data from two groups were statistically analyzed using two-tailed Student *t* tests (parametric unpaired) or Mann-Whitney U tests (nonparametric unpaired). When multiple comparisons were made, one-way or two-way ANOVA followed by Tukey’s multiple comparisons test or Sidak’s multiple comparisons test was performed using GraphPad Prism. Data are shown as means ± SEM unless otherwise stated.

## Declarations

### Ethics approval and consent to participate

This study was approved by the Ethics Committee of Tongji Hospital. Written informed consents were signed by the subjects recruited in the study or by the immediate family members in accordance with the Declaration of Helsinki.

### Authors’ contributions

SY and RX designed the study, analyzed, interpreted the data and drafted the paper; XZ, KJ, YT and JF participated in acquiring the data; CC, HL and DWW designed the work and drafted the paper.

## Data Availability

all data referred to in the manuscript will be available once our manuscript accepted

## Acknowledgements

We thank colleagues in Dr. Wang’s group for various technical help and stimulating discussion during this investigation.

